# Determinants and propagation of velocity uncertainty in 2D phase-contrast MRI

**DOI:** 10.64898/2026.06.01.26353730

**Authors:** Ana E. Rodríguez-Soto, Eleanor L. Schuchardt, Hari K. Narayan, Beth F. Printz, Sanjeet Hegde, Susan R. Hopkins, Francisco Contijoch

## Abstract

**Purpose:** To quantify the contributions of signal-to-noise ratio (SNR) and velocity-to-encoding ratio (v/VENC) to velocity uncertainty in phase-contrast (PC) MRI and to develop a framework for in vivo voxel-wise uncertainty estimation.

**Methods:** Through-plane 2D PC-MRI of the ascending aorta was acquired using multiple velocity encodings (150, 200, 300 cm/s) and flip angles (0°, 5°, 15°, 20°) to vary v/VENC and SNR. Voxel-wise SNR and velocity uncertainty maps were generated using empirically calibrated phase-noise modeling. Phase-resolved subject-level analyses were performed to quantify the relative contributions of SNR and |v|/VENC to percent velocity uncertainty (%unc). Uncertainty was propagated to flow, stroke volume (SV), and cardiac output (CO).

**Results:** Velocity uncertainty varied substantially across the cardiac cycle and depended on both SNR and |v|/VENC. Across cardiac phases, |v|/VENC accounted for most explained variance in %unc (partial R^2^=0.666), while SNR provided a smaller but meaningful contribution (partial R^2^=0.287; full R^2^=0.909). Near peak systole, SNR contributed more strongly while overall uncertainty remained low. In contrast, diastolic %unc became unstable as velocity approached zero. These effects were most pronounced at low |v|/VENC, where higher VENC settings increased uncertainty despite similar SNR. SV uncertainty ranged from 0.27% to 1.07% across VENC×FA protocols.

**Conclusion:** Velocity uncertainty in PC-MRI depends on both SNR and VENC adequacy in a physiologically phase-dependent manner. Relative uncertainty may become inadequate for precise quantification in low-flow applications, such as diastolic regurgitant jets, despite adequate SNR. Spatiotemporal uncertainty mapping provides a framework for uncertainty-aware PC-MRI acquisition and interpretation.

## 1. Introduction

Phase-contrast magnetic resonance imaging (PC-MRI) is widely used for noninvasive quantification of blood flow and velocity in cardiovascular, cerebrovascular, and other vascular applications ^1–4^. Accurate and precise velocity measurements are critical for estimating volumetric flow, pressure gradients, and derived hemodynamic metrics ^2–4^. However, the precision of PC-derived, which is dependent on acquisition parameters and physiologic flow conditions, is not routinely quantified in clinical practice.

The precision of PC-MRI measurements is commonly described using the velocity-to-noise ratio (VNR), defined as the ratio between measured velocity and its standard deviation in static tissue (VNR=|v|/σ_v_) ^5^. Under ideal assumptions of Gaussian, independent complex noise, velocity noise scales inversely with signal-to-noise ratio (SNR) and proportionally with velocity encoding (VENC), yielding the approximate relationship ^5,6^:

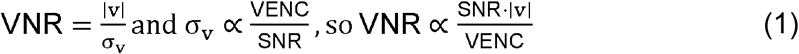

This expression emphasizes the two major determinants of measurement precision that can vary across scans: magnitude SNR and the ratio between velocity magnitude in the vessel of interest and VENC (|v|/VENC). While the role of SNR is widely appreciated, the influence of VENC adequacy has been less frequently emphasized. In practice, VENC is selected to avoid aliasing ^6^. However, conservative (higher) VENCs lead to lower |v|/VENC ratios and diminished velocity precision, particularly during low-flow phases of the cardiac cycle. This can be of high clinical significance in mixed stenotic and regurgitant lesions. The incorrect setting of VENC can decrease VNR, even when SNR is unchanged.

Despite these well-known relationships, the relative contributions of SNR and (|v|/VENC) to VNR are not well characterized. SNR varies across the field of view (FOV) and velocity magnitudes varying across cardiac phases, physiologic states, and between subjects ^2^. As a result, VNR will be spatially and temporally heterogeneous so a single global estimate of data quality may not adequately reflect the uncertainty of individual voxels or regions of interest ^7,8^. Therefore, there is a need for intuitive spatiotemporal maps of measurement reliability to better evaluate acquisition adequacy for clinical interpretation and consequential patient health management.

Lastly, VNR has limited clinical interpretability because its relationship to the precision of PC MRI derived flow measurements is not intuitive ^3,9^. Measurement uncertainty, in this case velocity uncertainty, has many potential benefits over the focus on VNR. First, it can be expressed directly in physical units (cm/s) or as percent uncertainty that can be used for direct analysis or error propagation ^9^. Second, it provides a more intuitive representation of measurement precision and allows spatially resolved assessment of uncertainty ^3^.

The purpose of this study was to develop spatiotemporal velocity uncertainty maps for 2D PC-MRI. To do so, we (i) measured voxel-wise SNR and velocity across cardiac phases, (ii) estimated the proportionality factor κ relating magnitude SNR to phase noise using stationary tissue measurements, (iii) quantified the independent contributions of 1/SNR and |v|/VENC to percent velocity uncertainty, and (v) generated spatially varying uncertainty maps to provide an intuitive framework for assessing acquisition quality and informing VENC selection.

## 2 Methods

### Subjects

This prospective study evaluated velocity uncertainty in 2D PC-MRI of the ascending aorta across multiple VENC and flip angle (FA) combinations. Twenty-five healthy volunteers underwent aortic 2D PC-MRI on a 1.5T scanner (GE Healthcare, Waukesha, WI, USA). All participants provided written informed consent in accordance with Institutional Review Board approval.

### Image Acquisition

All subjects underwent standard cardiac cine imaging for anatomical localization of the ascending aorta and aortic root prior to PC acquisition ^10^. Through-plane 2D PC-MRI was acquired in the proximal ascending aorta at the level of the sinotubular junction, with the imaging plane prescribed orthogonal to the aorta.

slice was performed perpendicular to the ascending aorta at the level of the aortic root ^2^.

For each subject, breath-held acquisitions were repeated across multiple VENC values (150, 200, 300 cm/s) and FA (0°, 5°, 15°, 20°) to systematically vary v/VENC and SNR. Imaging was performed using a retrospectively ECG-gated gradient-echo sequence with the following nominal parameters: repetition time (TR; 5.5, 5.6, and 5.5 ms for each VENC), echo time (TE; 3.3, 3.3, and 3.4 ms for each FA), field of view (FOV; 360×360 mm^2^), acquisition matrix (192×192), reconstruction matrix (256×256), in-plane spatial resolution (1.4×1.4 mm^2^), slice thickness (7 mm), receiver bandwidth (488 Hz/pixel), and 30 reconstructed cardiac phases. Vendor reconstructed magnitude and velocity images were used for all analyses.

### Regions of Interest

Ascending aorta regions of interest (ROIs) were manually delineated on magnitude images across all cardiac phases using OsiriX (Pixmeo SARL, Geneva, Switzerland) ^11^. ROIs were drawn frame-by-frame to account for cardiac translational motion and inter-scan positional differences.

Static tissue ROIs were automatically generated using the following algorithm. First, magnitude images were temporally averaged across the cardiac cycle and filtered with a 7×7 median kernel to reduce noise while preserving anatomical boundaries. Bias field inhomogeneity was corrected using the N4 algorithm ^12^ (SimpleITK implementation ^13^) with four-level multi-resolution optimization (100 iterations per level, convergence threshold 10^−5^) to normalize, as possible, intensities across the field of view. Zero-padded regions introduced by vendor reconstruction were automatically identified based on constant intensity values and excluded from all subsequent analyses.

A body mask was generated by separating tissue from background air using statistical comparison of local 7×7 neighborhoods to reference background regions sampled from image corners. Pixels were classified as tissue based on their z-score relative to the background intensity distribution (z>10). Morphological cleanup included hole filling, removal of connected components (<200 pixels), and a 3-pixel erosion to avoid boundary voxels. Background air regions were identified from non-zero voxels outside the body mask and used for noise estimation.

Regions exhibiting pulsatile flow were identified using temporal variation of the complex PC signal, *C*(*t*) *= M*(*t*)*e*^*iϕ*(*t*)^, where *M* is magnitude and *ϕ* is phase. For each pixel, the deviation from the temporal mean, Δ*C*(*t*) *= C*(*t*) - ⟨*C*(*t*)⟩_*t*_, was computed, and the maximum intensity projection of | Δ*C*(*t*) | across time was used to identify pixels with phase-varying flow. Vessel ROIs were detected using adaptive thresholding (Otsu, applied at the 60th percentile)^14^, followed by Gaussian smoothing (σ=0.8) and 5×5 median filtering. Isolated components (<10 pixels) were removed.

The final static tissue mask was defined as the intersection of the body mask with exclusion of the manually segmented aorta ROI and all detected vessel regions. A final 5-pixel erosion was applied to reduce partial volume effects at tissue boundaries.

### Signal-to-Noise Ratio Estimation

Magnitude SNR was computed voxel-wise for each cardiac phase as:

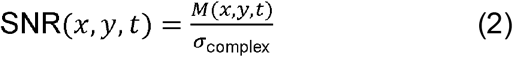

where *M*(*x, y, t*) denotes magnitude signal intensity and *σ*_complex_ represents the standard deviation of complex Gaussian noise.

Zero-padded regions (in the background) introduced by vendor reconstruction were excluded prior to noise estimation. Noise was estimated from valid (non-zero) air voxels using a median absolute deviation (MAD)-based estimator. Because magnitude noise in background regions follows a Rayleigh distribution ^15^, the measured magnitude noise was converted to equivalent complex Gaussian noise using the NEMA-recommended factor of 1.526 to obtain *σ*_complex_^16^.

To assess the stability of background-derived noise estimates, FA=0° acquisitions were used as a noise-only reference. Background air noise measured in FA≠0° acquisitions was spatially matched to corresponding FA=0° images, and agreement was evaluated using correlation and ratio analyses across acquisitions and cardiac phases. SNR maps were generated separately for each cardiac phase.

### Velocity Noise Model

To minimize systematic velocity offsets, a first-order polynomial background phase correction was applied using static tissue as reference ^17,18^. In PC-MRI, velocity is linearly related to the measured phase difference between velocity-encoded and reference acquisitions ^5,6,19^:

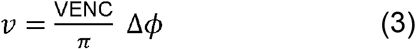

Using standard error propagation, velocity uncertainty is given by ^5,19^:

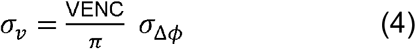

Thus, velocity precision is determined by the phase noise and can also be reported as a percent uncertainty. Normalizing by velocity removes the implicit scaling with VENC and expresses uncertainty in terms of SNR and *v*/VENC:

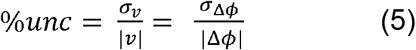

At sufficiently high SNR, the standard deviation of phase in a complex image with Gaussian noise is approximately inversely proportional to magnitude SNR (i.e., *σ*_*ϕ*_ ≈1/SNR) ^5,15^. Because velocity encoding involves subtraction of two independently acquired phase images (reference and velocity-encoded acquisitions), the variance of the phase difference is the sum of the individual variances. Under equal-SNR conditions:

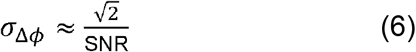

Substituting this relationship into the expression for *σ*_*ϕ*_ (Equation 4) yields the first-order proportionality:

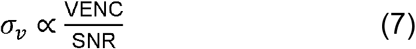

To express this relationship quantitatively, a proportionality factor κ has been used, where κ is a dimensionless factor relating magnitude SNR to phase-difference noise ^5,6,19^:

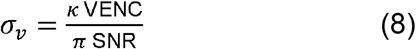

Under ideal Gaussian assumptions with independent reference and velocity-encoded acquisitions, subtraction of two phase images increases variance by a factor of two, yielding κ = √2^5,6,19^. However, *in vivo* deviations from ideal noise behavior and reconstruction-related correlations may alter this relationship ^9^, motivating empirical estimation of κ. Accurate estimation of σ_complex_ is required to ensure that SNR, and consequently κ and velocity uncertainty, are correctly scaled across the field of view.

### Empirical Estimation and Modeling of κ

The theoretical phase noise model assumes ideal Gaussian noise (σ_φ_=1/SNR) ^5,15^. In practice, reconstruction and processing steps introduce correlated noise that deviates from this behavior ^20,21^. The proportionality factor κ was therefore estimated empirically ^5,9^. κ was estimated voxel-wise in stationary tissue, where the true velocity is zero and any measured velocity reflects noise ^9,17^. The temporal standard deviation of velocity σ_v,static_ was computed and used to estimate κ as:

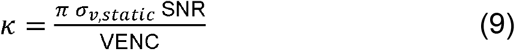

Where *σ*_*v*, static_ is the temporal standard deviation of velocity in static tissue and SNR was computed as described above.

### Estimation of Uncertainty Maps

Voxel-wise velocity uncertainty was computed as:

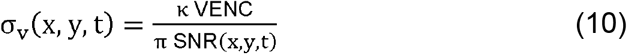

where κ was estimated empirically as described above. Percent velocity uncertainty was defined as:

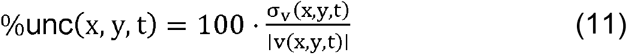

Spatially resolved uncertainty maps were generated for all subjects and acquisition protocols.

### Determinants of Percent Velocity Uncertainty

To quantify the independent contributions of SNR and |v|/VENC to percent uncertainty, two complementary analyses were performed. Median values of %unc, SNR, and |v|/VENC were computed within the aortic ROI for each subject and acquisition protocol (one observation per subject×protocol×cardiac phase). Partial R^2^ analysis was then performed, with percent uncertainty as the dependent variable. Phase-resolved analyses were performed across the cardiac cycle to quantify the relative contributions of SNR and |v|/VENC as a function of cardiac phase.

### Flow Rate and Stroke Volume Uncertainty Propagation

Aortic flow rate (Q) was computed at each cardiac phase by adding the voxel-wise velocities within the ascending aorta ROI and multiplying by voxel area ^2,3^. Flow rate uncertainty was propagated from voxel-wise velocity uncertainty maps. To account for spatial correlation of velocity noise, an effective-sample framework was applied in which the independent root-sum-of-squares (RSS) estimate was scaled by √(N/N_eff_), where N is the total number of voxels and N_eff_ is the effective number of independent voxels ^9,22^.

Stroke volume (SV) was computed by integrating flow across the cardiac cycle ^23^. Its uncertainty was estimated by propagating flow uncertainty across cardiac phases using an effective number of independent time points to account for temporal correlation ^9^. Although background noise exhibited minimal temporal correlation, an effective number of independent cardiac phases was estimated to account for residual physiological and reconstruction-related correlations in flow signals. For reference, SV was also computed under an independence assumption using RSS propagation across cardiac phases.

Cardiac output (CO) was calculated as CO=SV×HR, where heart rate (HR) was obtained from DICOM header information ^2^.

### Statistical Analysis

All statistical analyses were performed using Python (3.10) with standard libraries (numpy 2.2, scipy 1.15, statsmodels 0.14, scikit-learn 1.7). Continuous variables are reported as median (interquartile range, IQR) unless otherwise specified.

For noise validation, agreement between background-derived noise estimates from FA≠0° acquisitions and corresponding FA=0° noise measurements was assessed using Pearson correlation coefficients at both the acquisition level (one value per subject×VENC×FA) and the cardiac phase level (frame-by-frame comparisons across all cardiac phases). The ratio between FA≠0° background air noise and FA=0° noise was computed to assess consistency of noise scaling across protocols.

As an additional quality-control analysis, the standard deviation of magnitude signal within the aorta ROI in FA=0° acquisitions was compared to background-derived σ_complex_. Their ratio was also evaluated to assess agreement of noise magnitude within the FOV.

Temporal stability of background noise estimates within each acquisition was evaluated using Pearson correlation between FA=0° and FA≠0° background frame-mean time series across cardiac phases. Differences in κ across VENC and flip angle (FA) were assessed using nonparametric Kruskal-Wallis tests.

For determinant analysis, values of percent velocity uncertainty (%unc), SNR, and normalized velocity (|v|/VENC) were computed as the median within the aortic ROI at each cardiac phase, yielding one observation per subject×VENC×FA×cardiac phase. Linear models were used to estimate the relative contributions of predictors to %unc. Partial R^2^ was computed as the change in model R^2^ upon removal of each predictor. A two-sided p-value < 0.05 was considered statistically significant.

## 3. Results

### Subjects and acquisitions

Twenty-five healthy volunteers (12 male, 13 female) were recruited through institutional advertisement and word of mouth. Inclusion criteria included age ≥18 years and ability to safely undergo MRI. Exclusion criteria included known cardiovascular disease, contraindications to MRI, and pregnancy. Participants had a median age of 30 years (IQR 29), height 172.7 cm (15.2 cm), median weight 68.0 kg (17.7 kg), and body mass index (BMI) of 24.4 kg/m^2^ (IQR 4.7 kg/m^2^). Self-reported race included White (n=17), Asian (n=4), Black (n=1), more than one race (n=2), and unknown (n=1). Ethnicity was reported as Hispanic or Latino (n=8) and Not Hispanic or Latino (n=17).

All participants completed ascending aorta 2D PC-MRI across repeated combinations of VENC and FA. Heart rate during PC acquisitions was 62.4±8.9 bpm (range 45-96 bpm). Peak systolic aortic velocity across subjects was 56.5±16.8 cm/s (range 20-96). Across all aortic ROI voxels and cardiac phases, the distribution of |v|/VENC spanned 0.01 to 0.83, with higher |v|/VENC observed at lower VENC settings and during systole (**Table 1**).

**Table 1.** Aortic v/VENC, SNR and velocity uncertainty (%unc) at peak systole and diastole. Values are median (IQR, range).

| VENC<br>(cm/s) | FA<br>(°) | \|v\|/VENC peak<br>systole | \|v\|/VENC<br>diastole | SNR systole | SNR diastole | %unc<br>systole | %unc<br>diastole |
| --- | --- | --- | --- | --- | --- | --- | --- |
| 150 | 5 | 0.43<br>(0.16, 0.25-0.80) | 0.03<br>(0.02, 0.02-0.05) | 15.5<br>(13.5, 7.1-28.3) | 14.5<br>(12.7, 7.2-27.4) | 5.9<br>(5.0, 0-17.8) | 90.8<br>(32.5, 0-134) |
|  | 15 | 0.43<br>(0.15, 0.23-0.82) | 0.02<br>(0.01, 0.01-0.03) | 40.0<br>(29.5, 17.1-80.0) | 26.9<br>(14.7, 13.2-50.2) | 2.0<br>(1.9, 0-10.7) | 71.4<br>(36.1, 0-127) |
|  | 20 | 0.44<br>(0.14, 0.25-0.82) | 0.02<br>(0.01, 0.01-0.04) | 49.6<br>(37.5, 22.4-97.9) | 28.0<br>(13.4, 12.7-45.1) | 1.9<br>(1.6, 0-5.2) | 71.5<br>(28.6, 0-117) |
| 200 | 5 | 0.30<br>(0.11, 0.17-0.62) | 0.03<br>(0.02, 0.02-0.06) | 16.1<br>(12.1, 5.4-50.4) | 15.9<br>(10.7, 5.2-49.9) | 10.5<br>(6.4, 2.5-57.8) | 93.7<br>(36.2, 62-154) |
|  | 15 | 0.32<br>(0.13, 0.17-0.62) | 0.02<br>(0.01, 0.01-0.03) | 44.4<br>(35.6, 14.8-126) | 31.2<br>(20.8, 11.0-82.2) | 2.9<br>(1.9, 0.9-15.8) | 78.3<br>(42.7, 41-115) |
|  | 20 | 0.32<br>(0.12, 0.05-0.60) | 0.02<br>(0.0, 0.01-0.03) | 43.1<br>(36.3, 25.3-153) | 30.6<br>(21.2, 14.3-80.9) | 2.5<br>(2.1, 0.8-16.8) | 71.1<br>(26.3, 31-105) |
| 300 | 5 | 0.21<br>(0.07, 0.11-0.40) | 0.02<br>(0.01, 0.02-0.05) | 16.1<br>(12.6, 6.9-28.7) | 15.7<br>(12.6, 7.3-29.6) | 14.2<br>(9.1, 3.6-38.8) | 88.8<br>(39.1, 70-145) |
|  | 15 | 0.22<br>(0.07, 0.12-0.42) | 0.01<br>(0.01, 0.01-0.03) | 39.2<br>(27.0, 18.9-78.2) | 32.5 (<br>(22.7, 14.0-52.8) | 4.5<br>(3.6, 1.3-20.4) | 77.6<br>(34.5, 47-134) |
|  | 20 | 0.23<br>(0.07, 0.12-0.42) | 0.02<br>(0.01, 0.01-0.03) | 41.9<br>(31.3, 23.3-105) | 28.3<br>(19.3, 13.9-70.3) | 3.8<br>(2.9, 1.1-10.6) | 73.0<br>(34.1, 40-123) |

Aortic ROI cross-sectional area, quantified as the spatial correlation footprint, was consistent across subjects. Across all 25 participants at VENC150/FA20°, 18 of 25 subjects (72%) had an area of 19.6 voxels, with 6 subjects having a smaller area of 7.1 voxels and 1 subject having a larger area of 38.5 voxels. When summed across all VENC×FA protocols per subject, the total voxel contribution per participant had a median of 157.9 voxels (IQR 126.4-176.7; range 101.3-220.7), with a coefficient of variation of 19.3%, indicating broadly comparable contributions across subjects with no single participant dominating the pooled analysis.

### Aortic v/VENC, SNR and percent uncertainty

SNR increased with FA and was consistently higher in systole than diastole across all VENC settings (**Table 1**). For example, at VENC 150 cm/s, median systolic SNR increased from 15.5 at FA 5° to 49.6 at FA 20°, with similar trends observed across VENC 200 and 300 cm/s.

Percent velocity uncertainty (%unc) displayed dependence on both SNR and |v|/VENC. At peak systole, %unc decreased with increasing FA and decreasing VENC, ranging from 14.2% (IQR 9.1) at VENC 300/FA 5° to 1.9% (1.6) at VENC 150/FA 20°. In contrast, diastolic %unc remained high (range 71.1-93.7%) across all protocols due to low |v|/VENC, with median values ranging from 71-94% despite SNR levels similar to peak systole.

Temporal variation in |v|/VENC across the cardiac cycle is shown in **Figure 1**. Peak systolic |v|/VENC occurred consistently near phase 5 across all VENC settings, with lower VENC yielding higher |v|/VENC throughout systole. Diastolic values remained near zero across all protocols, consistent with the elevated uncertainty shown in **Table 1**.

**Figure 1.**
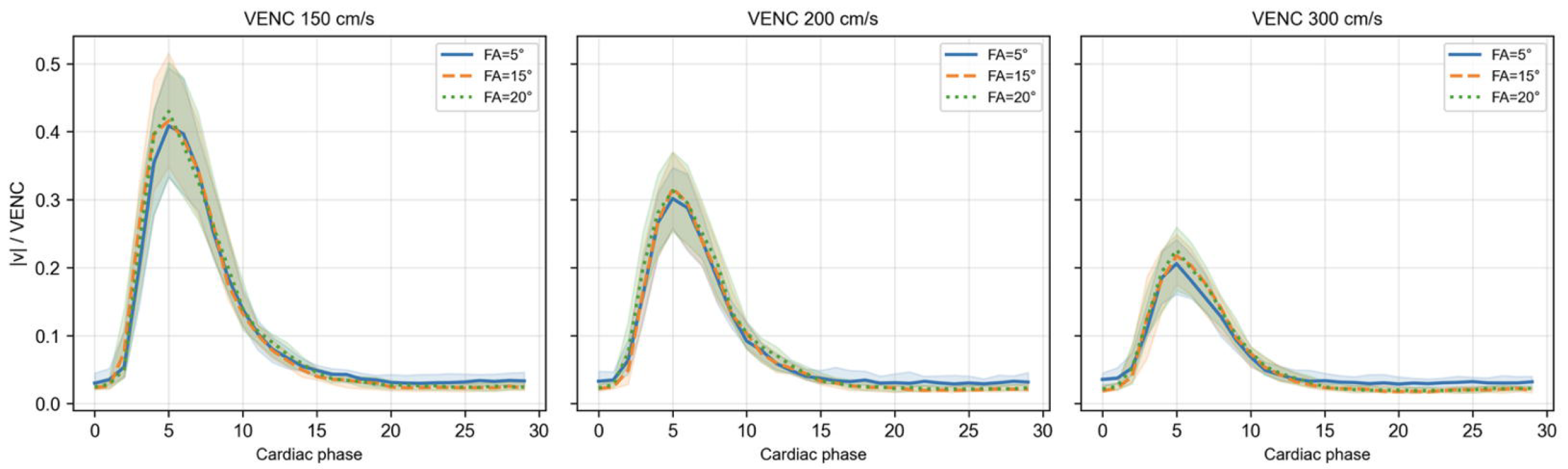
Temporal variation of |v|/VENC across the cardiac cycle. Median |v|/VENC across the cardiac cycle is shown for VENC=150, 200, and 300 cm/s. Curves correspond to FAs (5°, 15°, 20°), and shaded regions represent interquartile range across subjects. Lower VENC yield higher |v|/VENC throughout systole, while diastolic values remain near zero across all protocols. |v|/VENC is largely consistent across FAs, with minor differences reflecting SNR-driven variability.

### Validation of noise estimates

Background-derived σ_complex_ estimates demonstrated good agreement with corresponding FA=0° reference acquisitions across subjects and protocols (**Supplementary Figure S1**). Noise estimates showed consistent scaling across acquisition conditions with minimal temporal correlation across cardiac phases, supporting the use of background-derived noise estimation for voxel-wise SNR and uncertainty mapping.

### Empirical estimation of κ

The empirically estimated κ across subjects was 1.90 (IQR: 1.49-2.05), compared to the theoretical value of √2, a ~35% increase relative to the ideal Gaussian noise assumption. κ showed no significant variation across VENC (p=0.70) or flip angle (p=0.78).

### Relative contributions of SNR and VENC adequacy to uncertainty

Using phase-resolved subject-level observations of %unc, normalized velocity (|v|/VENC) was the dominant determinant, accounting for the majority of explained variance across the cardiac cycle (partial R^2^=0.666), while SNR contributed a smaller but meaningful fraction (partial R^2^=0.287; **Figure 2**). The combined model explained most of the observed variation in %unc (full R^2^=0.909).

**Figure 2.**
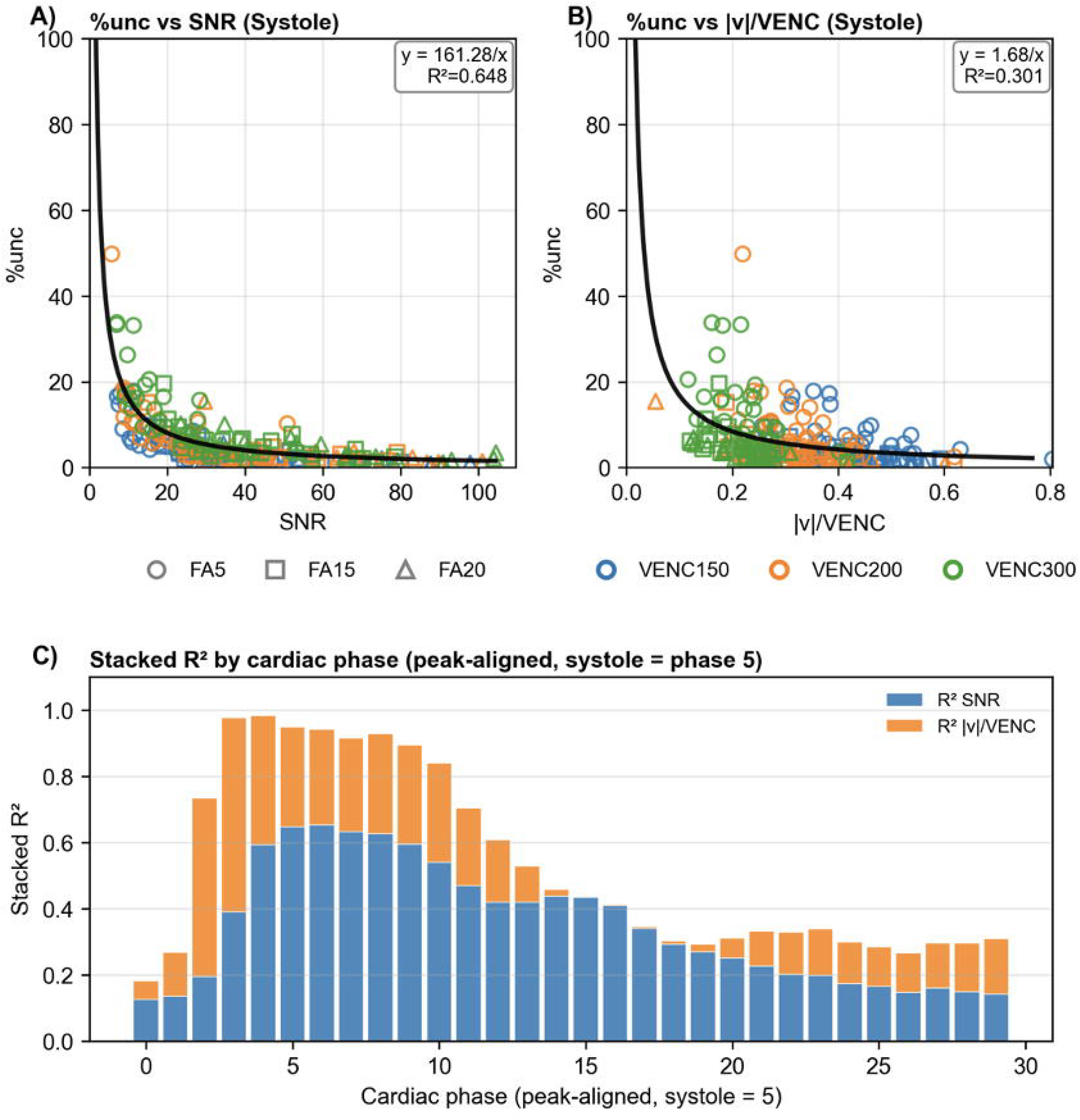
Effect of VENC on velocity uncertainty at fixed flip angle (FA=20°) during peak systole. Representative magnitude (MAG), velocity (VEL), signal-to-noise ratio (SNR), velocity uncertainty (σ_v_), and percent uncertainty (%unc) maps are shown for VENC=150 (top row), 200 (middle row), and 300 cm/s (bottom row) in a single subject. SNR (middle column) is comparable across VENC settings at fixed FA, while σ_v_ increases (fourth column) with VENC, consistent with σ_v_ ∝ VENC/SNR. As a result, %unc increases (last column) with VENC due to reduced |v|/VENC, despite similar SNR. Within the aortic ROI, %unc remains lowest during systole when velocity is high and increases in surrounding regions with lower SNR and near-zero velocity.

During systolic phases, %unc demonstrated inverse relationships with both SNR and normalized velocity (|v|/VENC) (**Figure 2**). The relationship with SNR followed y=161.28/SNR (R^2^=0.648), while the relationship with |v|/VENC followed y=1.68/(|v|/VENC) (R^2^=0.301). Although both predictors influenced uncertainty, phase-resolved analysis demonstrated that their relative contributions varied substantially across the cardiac cycle (**Figure 2C**). Near peak systole, SNR accounted for a larger fraction of explained variance while overall uncertainty remained comparatively low. In contrast, during low-flow phases, |v|/VENC became the dominant determinant of %unc, reflecting the divergence of relative uncertainty as velocity approached zero.

These relationships were consistent across subjects and acquisition protocols. Lower VENC and higher FA were associated with reduced %unc across velocity ranges, with protocol-specific relationships shown in **Figure S3**. Across protocols, the strongest velocity-%unc relationships were observed at lower VENC and higher FA combinations, consistent with improved precision under conditions of higher SNR and higher |v|/VENC.

### Spatially resolved uncertainty maps across VENC and FA

Consistent with the determinant analysis, voxel-wise uncertainty maps demonstrated spatial heterogeneity across 2D PC protocols (**Figures 3 and 4**). At peak systole, where velocity magnitude is high, spatial patterns of %unc were primarily governed by acquisition parameters rather than temporal variation. At fixed flip angle (FA=20°), SNR was comparable across VENC settings, while velocity uncertainty (σ_v_) increased with VENC, consistent with σ_v_ ∝ VENC/SNR (**Figure 3**). As a result, %unc increased with VENC due to reduced |v|/VENC, despite similar SNR. At fixed VENC (150 cm/s), increasing FA led to higher SNR and corresponding reductions in σ_v_, consistent with σ_v_ ∝ 1/SNR (**Figure 4**). Consequently, %unc decreased with increasing FA. Across all protocols, spatial patterns of %unc remained strongly dependent on velocity magnitude, with lowest uncertainty within the aortic lumen during systole and elevated uncertainty in surrounding regions characterized by low velocity and/or low SNR.

**Figure 3.**
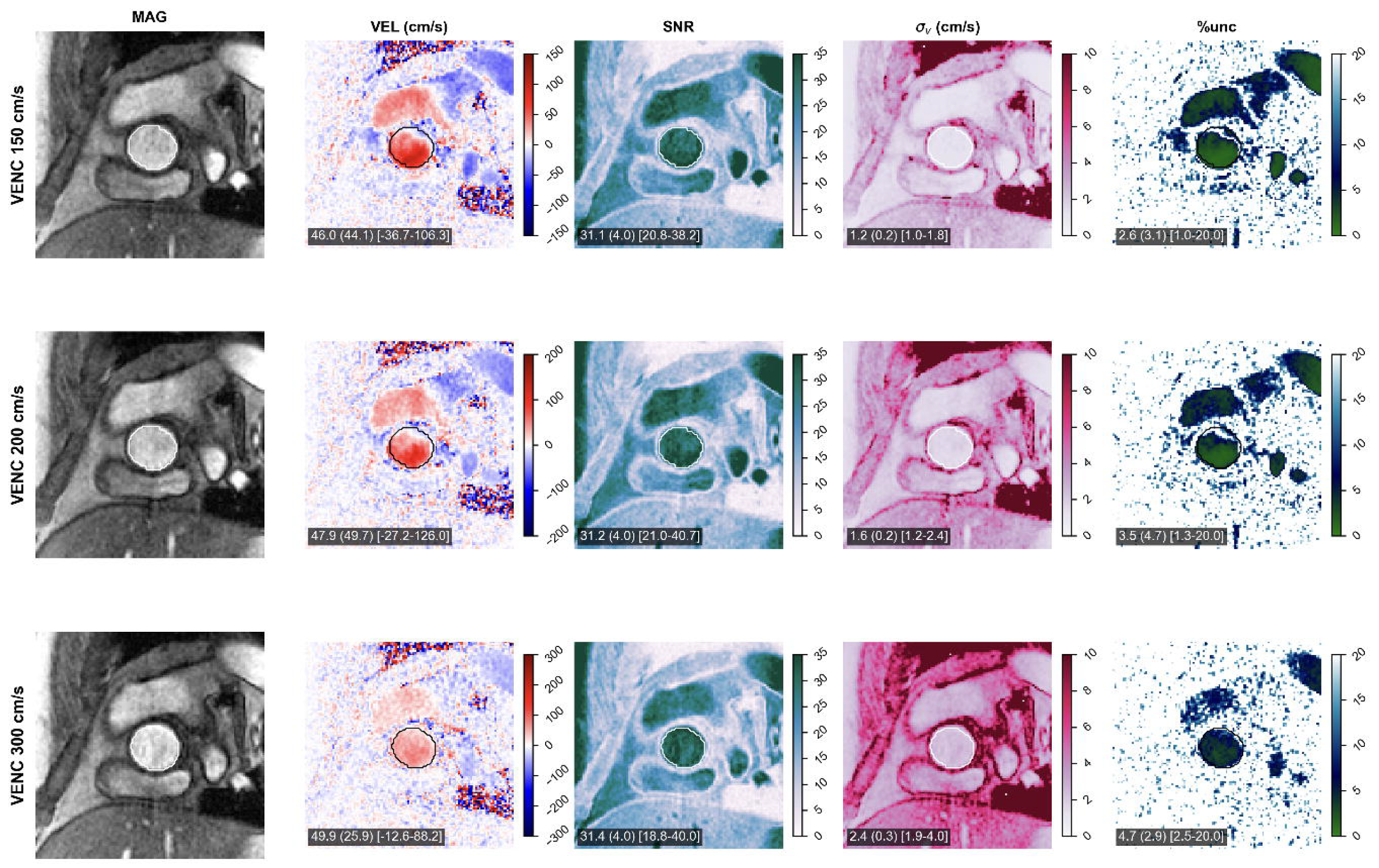
Effect of flip angle on velocity uncertainty at fixed VENC (150 cm/s) during peak systole. Representative MAG, VEL, SNR, σ□, and %unc maps are shown for FA=5° (top row), 15° (middle row), and 20° (bottom row) in a single subject. Increasing FA leads to higher SNR (middle column), resulting in reduced σ□ (fourth column), consistent with σ□ ∝ 1/SNR. Consequently, %unc decreases (fifth column) with increasing FA at fixed VENC. Spatial patterns of %unc remain strongly dependent on velocity magnitude, with lowest uncertainty in high-velocity regions and elevated uncertainty in regions of low velocity or low SNR.

**Figure 4.**
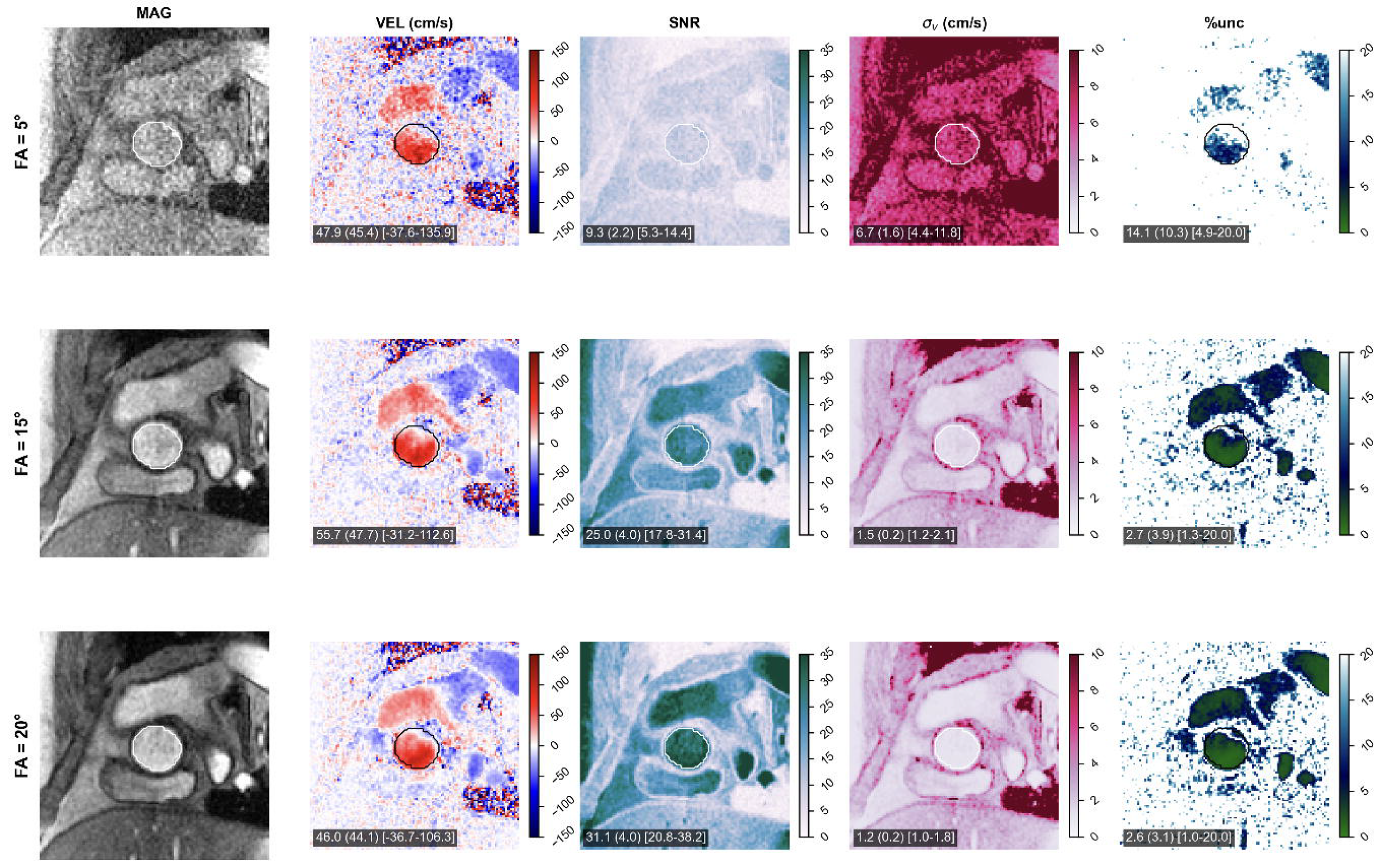
Effect of flip angle on velocity uncertainty at fixed VENC (150 cm/s) during peak systole. Representative MAG, VEL, SNR, σ□, and %unc maps are shown for FA=5°, 15°, and 20° in a single subject. Increasing FA leads to higher SNR, resulting in reduced σ□, consistent with σ□ ∝ 1/SNR. Consequently, %unc decreases with increasing FA at fixed VENC. Spatial patterns of %unc remain strongly dependent on velocity magnitude, with lowest uncertainty in high-velocity regions and elevated uncertainty in regions of low velocity or low SNR.

### Flow uncertainty with spatial correlation correction

Uncertainty propagation from voxel-wise σ□ maps to instantaneous flow demonstrated that spatial correlation of velocity noise substantially inflates ROI-level uncertainty relative to the independent-voxel RSS assumption (**Figure 5**). This correction used the effective-sample framework described in Methods ^9,22^, in which spatial correlation length estimated from stationary tissue was used to reduce the apparent number of independent voxels within the aortic ROI. Spatial autocorrelation functions computed from stationary tissue were well-described by an exponential decay model, yielding a spatial correlation length L_spatial_ of 2.5 mm across subjects. This corresponded to an effective number of independent voxels N_eff_ that was markedly smaller than the total number of aortic ROI voxels (median reduction factor 0.06; median ROI size 343 voxels, IQR 270-384).

**Figure 5.**
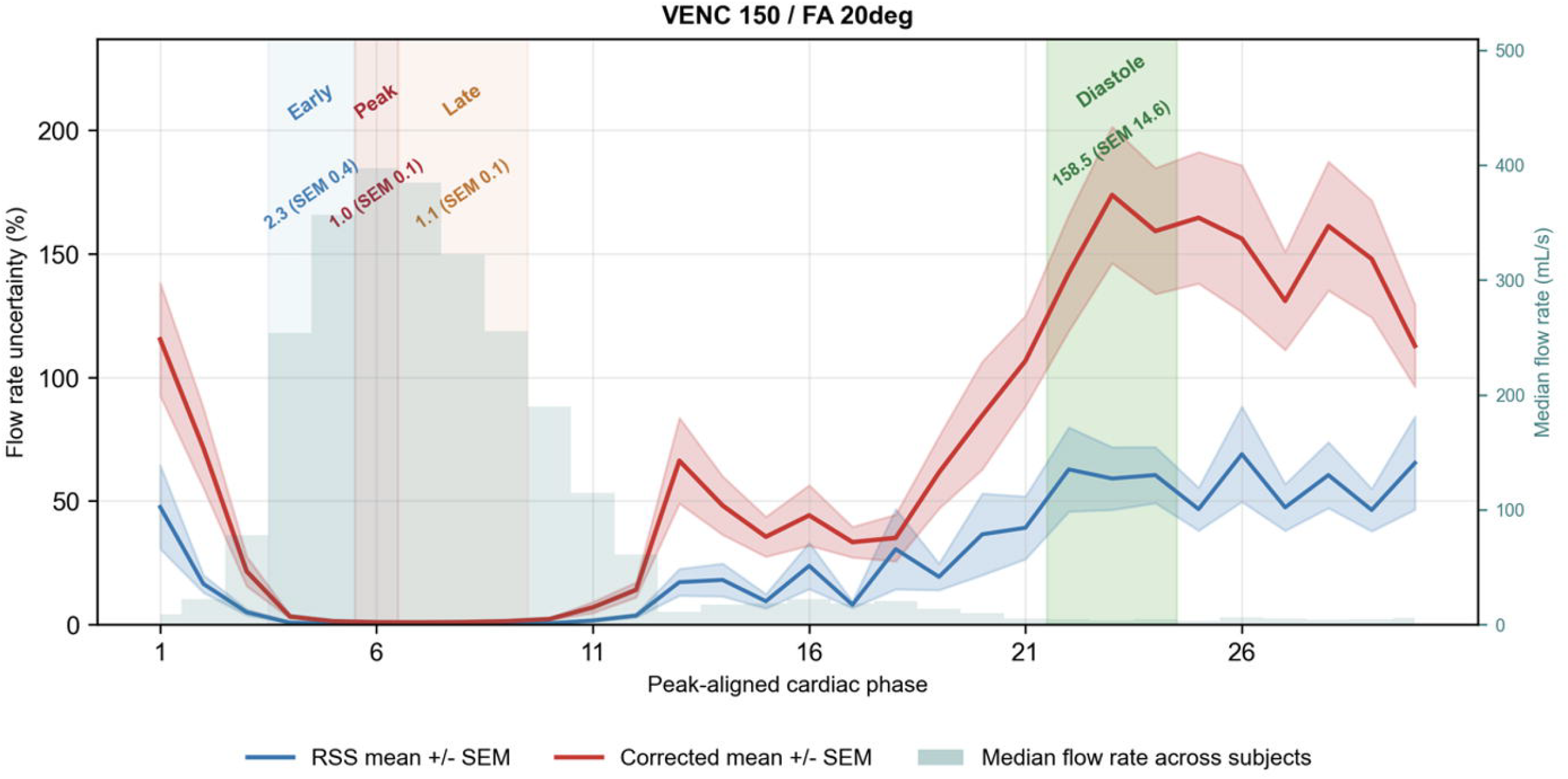
Effect of spatial correlation on phase-resolved flow uncertainty. Phase-aligned percent flow uncertainty across the cardiac cycle for VENC 150 cm/s and FA 20°, comparing independent-voxel RSS estimates and spatially corrected estimates after effective-sample correction. The median flow-rate curve across subjects is overlaid to illustrate the relationship between flow magnitude and percent uncertainty. Shaded regions indicate early systole, peak systole, and late systole. Spatial correlation leads to substantially higher estimated uncertainty compared to the independent-voxel assumption, with the largest relative differences observed during phases of higher flow.

After applying the effective-sample correction, instantaneous flow uncertainty increased by a factor of 4.4 relative to the independent RSS estimate. Across all protocols, median instantaneous flow uncertainty was 9.2 mL/s (IQR: 5.8–14.3), corresponding to a percent uncertainty of 60.0% (IQR: 40.0–89.0%).

Diastolic phases were excluded from summary statistics due to instability arising from near-zero flow, where percent uncertainty becomes ill-conditioned by definition. To account for this behavior, cardiac phases were grouped into physiologically meaningful bins based on peak-aligned flow: early systole, peak systole, and late systole (**Figure 5**). Spatially corrected percent uncertainty was lowest near peak systole, where flow magnitude was highest, and increased during early and late systole. Phase-binned analysis demonstrated that percent uncertainty was consistently lowest during peak systole and increased during early and late systole across all VENC and FA combinations (**Figure S2**). For example, at VENC 150 cm/s and FA 20°, mean percent uncertainty was approximately 2.3% in early systole, 1.0% at peak systole, and 1.1% in late systole, compared to substantially higher and inherently unstable values during diastole.

Similar trends were observed across all protocols, with higher VENC and lower flip angle associated with increased percent uncertainty. This phase-based reporting avoids bias introduced by variable exclusion of low-flow frames and enables consistent comparison across subjects and acquisition protocols.

### Stroke volume uncertainty with temporal correlation correction

The uncertainty of SV was computed by propagating instantaneous flow uncertainty across retrospectively gated cardiac phases. Under the independent-frame assumption, median SV uncertainty across protocols was 0.38 mL (IQR: 0.26-0.59), corresponding to a percent uncertainty of 0.54% (IQR: 0.4-0.8%). Accounting for temporal correlation resulted in only a minimal change in SV uncertainty, indicating that temporal dependence between cardiac phases was negligible. Despite the substantial increase in instantaneous flow uncertainty after spatial correlation correction, its impact on integrated metrics was minimal. Cardiac output (CO) uncertainty was 0.027 L/min (~0.5%).

SV and CO uncertainty increased significantly with VENC (p<0.001) and with lower flip angle (p<0.001). Median SV and CO uncertainty was 0.38% (IQR: 0.24-0.53%) at VENC150 cm/s, 0.52% (0.39-0.71%) at VENC 200 cm/s, and 0.84% (0.56-1.18%) at VENC 300 cm/s. By FA, median uncertainty was 0.80% (0.52-1.12%) at FA 5°, 0.50% (0.32-0.69%) at FA 15°, and 0.47% (0.29-0.63%) at FA 20°. The VENC×FA interaction was not significant (p=0.983), indicating that both factors contributed independently and additively to integrated flow uncertainty.

## 4. Discussion and Conclusions

In this study, we developed and evaluated a framework for quantifying spatiotemporal velocity uncertainty in 2D PC-MRI. By combining voxel-wise SNR estimation, empirical calibration of phase noise (κ), and propagation of uncertainty to flow-derived metrics, we show that velocity precision is heterogeneous across both space and time and is governed by a combination of acquisition parameters and physiologic flow conditions.

The velocity to VENC ratio (|v|/VENC) is the dominant determinant of percent velocity uncertainty, while SNR provides a secondary but still meaningful contribution. This relationship reflects the nonlinear dependence of percent uncertainty on velocity magnitude, while uncertainty increases rapidly as |v| approaches zero. As a result, even in the presence of adequate SNR, low-velocity regions, particularly during diastole, have elevated uncertainty. Our findings highlight that VENC selection plays a critical role in determining measurement precision, beyond its conventional use for avoiding velocity aliasing ^2,3,5^.

Previous studies have demonstrated that suboptimal VENC selection reduces VNR and increases measurement error, particularly in low-velocity regimes, while increases in SNR provide more modest improvements once a sufficient baseline is achieved. However, these relationships have primarily been described using global or ROI-based metrics ^2,7–9^. This work extends the VNR framework by providing voxel-wise, phase-resolved quantification of uncertainty and decomposing the relative contributions of SNR and normalized velocity in vivo ^9^. Prior work has also explored variable or adaptive VENC strategies, in which lower VENC values are used during low-flow phases and higher VENC values during systole to improve velocity sensitivity and vascular conspicuity ^24,25^. These approaches were largely motivated empirically. The present results provide a quantitative framework explaining these observations by demonstrating that uncertainty depends strongly on both physiologic flow state and |v|/VENC throughout the cardiac cycle.

Our results demonstrate that selection of VENC to maximize |v|/VENC has a larger impact on velocity precision than further increases in SNR once moderate SNR is achieved. In practical terms, excessively high VENC settings reduce velocity sensitivity and increase uncertainty, even when SNR is preserved. Conversely, optimization of SNR through higher FA or increasing averages provides incremental improvements in precision but does not compensate for suboptimal VENC selection. This implies that improving VENC selection is often more effective than increasing SNR for reducing velocity uncertainty ^2,5,6^. Further, the relative contributions of SNR and |v|/VENC varied substantially across the cardiac cycle. Around peak systole, where velocities are highest and relative uncertainty remains low, SNR contributed more strongly to the observed variance in %unc. During lower-flow phases, the explanatory power of both predictors decreased substantially as velocity approached zero and percent uncertainty became increasingly ill-conditioned. These findings suggest that optimal VENC selection may depend on the specific physiologic flow pattern or clinical application of interest, rather than solely on avoidance of aliasing.

Although magnitude SNR is often treated as an acquisition-dependent quantity, our data demonstrated cardiac phase-dependent SNR variation within the aortic ROI. This likely reflects flow-related changes in blood magnitude signal, including inflow-related refresh of spins, saturation during slower flow, intravoxel dephasing, and partial-volume effects across the cardiac cycle. Thus, uncertainty in PC-MRI is influenced not only by prescribed acquisition parameters such as FA and VENC, but also by physiologic flow conditions that alter both velocity magnitude and effective SNR.

Empirical estimation of the proportionality factor κ revealed a consistent deviation from the theoretical value of √2 ^5^, with an approximately 35% increase observed across subjects. This discrepancy likely reflects non-ideal noise behavior arising from reconstruction processes, coil sensitivity variations, and other sources of noise in vivo. These factors introduce dependencies between velocity-encoded and reference acquisitions that are not captured by ideal Gaussian assumptions ^15,20,21^. The stability of κ across VENC and FA combinations supports its use as a global scaling factor for velocity uncertainty estimation, while emphasizing the importance of empirical calibration when applying noise models to in vivo data ^5,15,20,21^.

These findings also support the use of a global noise estimate (σ_complex_) for voxel-wise SNR mapping. While this approach assumes spatially uniform noise after exclusion of zero-padded regions, residual spatial variation may persist due to coil sensitivity and reconstruction effects ^20,21^. Despite this, the consistency of noise estimates across acquisition conditions suggests that a global noise model provides a reasonable approximation for uncertainty estimation.

We also demonstrate that spatial correlation of velocity noise inflates ROI-level uncertainty relative to independent-voxel assumptions. Incorporation of an effective-sample framework ^9,22^ increased instantaneous flow uncertainty by a factor of approximately 4.4, indicating that conventional RSS propagation underestimates uncertainty when spatial correlations are ignored ^9,22^. Despite this increase at the voxel and instantaneous flow level, propagated uncertainty in integrated metrics such as SV and CO remained low due to temporal averaging across the cardiac cycle. In contrast, temporal correlation between cardiac phases was minimal, supporting the assumption of independence in temporal uncertainty propagation ^9^.

Temporal variation of uncertainty was strongly dependent on the cardiac cycle. Phase-resolved analysis demonstrated that uncertainty is minimized near peak systole, where both velocity magnitude and SNR are highest, and increases during early and late systole, with markedly elevated and inherently unstable values during diastole due to near-zero flow. This instability arises from the definition of percent uncertainty, which becomes ill-conditioned as flow approaches zero. Because uncertainty is inherently time-dependent, reporting of flow-derived metrics should account for physiologic phase to avoid bias introduced by low-flow conditions^9,26^.

This work has several implications for clinical and research applications. First, voxel-wise uncertainty maps provide an intuitive and spatially resolved assessment of measurement precision, enabling identification of regions where quantitative interpretation may be limited ^9^. Second, phase-resolved uncertainty analysis offers a principled approach for selecting cardiac phases for quantitative flow assessment, favoring time points with higher velocity magnitude and lower relative uncertainty ^2,3^. Third, these results support the use of uncertainty-aware acquisition strategies, in which VENC is selected based on expected physiologic velocities to maximize |v|/VENC and minimize measurement uncertainty ^2,3,9^. Together, these approaches provide a framework for improving both the reliability and interpretability of quantitative PC-MRI measurements. These findings are relevant for low-flow applications such as venous flow imaging, regurgitant flow quantification, collateral flow assessment in congenital heart disease, cerebrospinal fluid flow measurements, and imaging of smaller vessels, where conservative high-VENC prescriptions may substantially increase uncertainty despite adequate magnitude SNR.

This study has several limitations. First, the analysis was performed in healthy volunteers using a single-vendor 1.5T 2D PC-MRI implementation and reconstruction, and uncertainty behavior may differ across pipelines, field strengths, and clinical populations. Second, the proposed framework was evaluated in the ascending aorta and may not directly generalize to vessels with substantially different flow characteristics, including turbulent or highly complex flow patterns. Third, this study focuses on quantifying measurement precision and uncertainty rather than accuracy, which is determined by several factors including spatial and temporal resolution, background phase offsets, flow complexity, and reconstruction-related biases that were not directly evaluated in this study ^2,4^.

Third, estimation of voxel-wise SNR was based on a global background-derived noise model using σ_complex_ measured from air regions outside the body. This approach assumes spatially uniform noise throughout the field of view after reconstruction. In practice, spatial variation in noise may persist due to coil sensitivity profiles, parallel imaging reconstruction, filtering, and other vendor-specific processing steps ^20,21^. As a result, local SNR and uncertainty estimates may not fully reflect spatially varying noise amplification, particularly near the periphery of the field of view or in regions with lower coil sensitivity. Future work will focus on evaluating this framework in clinical populations and vessels with more complex and pathologic flow patterns.

Finally, while this study focuses on 2D PC-MRI, these findings also have implications for 4D flow MRI. In 4D flow acquisitions, additional sources of noise, including undersampling, trajectory errors, and reconstruction regularization ^26–28^, are present and may further increase velocity uncertainty and introduce additional spatial and temporal correlations. Extension of this uncertainty framework to 4D flow MRI will therefore require dedicated modeling of these effects and may benefit from adaptive acquisition strategies that jointly optimize SNR and VENC. More broadly, the observed dependence of uncertainty on physiologic flow regime supports the development of physiology-aware or adaptive VENC strategies rather than a one-size-fits-all approach designed solely to avoid aliasing.

## Data Availability

All data produced in the present study are available upon reasonable request to the authors

## DATA AVAILABILITY STATEMENT

The data that support the findings of this study are available from the corresponding author upon reasonable request, subject to institutional and ethical approval requirements.

## ACKNOWLEDGEMENTS

National Institute of Health, National Heart Lung and Blood Institute grants R56HL159710, R01HL162671., and NHLBI K23HL163456.

