## Supplemental Material for "Determinants and propagation of velocity uncertainty in 2D phase-contrast MRI"

**SUPPLEMENTARY MATERIAL**

Across subjects and protocols, σ_complex_ measured from background air in FA≠0° acquisitions correlated with that measured from FA=0° acquisitions at both the protocol level (r=0.606, p<0.001) and cardiac-phase level (r=0.761, p<0.001; **Figure S1A-B**). The ratio σ_air_/σ_FA0_ was 1.00 (IQR: 1.00-1.05), indicating consistent noise scaling across acquisition conditions. Within-protocol temporal stability, quantified as Pearson r^2^ between FA=0° and FA≠0° background frame-mean time series, was low (median r^2^=0.033, IQR: 0.008-0.094; **Figure S1C**), consistent with random frame-to-frame noise variation. FA=0° aorta ROI measurements were used as an additional quality-control assessment, comparing the standard deviation of magnitude signal within the aorta to the background-derived σ_complex_ (**Figure S1D**). The ratio between aortic signal standard deviation and background-derived σ_complex_ was 0.66 (median), with limited correlation (r=0.334), indicating the presence of low but nonzero signal within the aorta ROI.

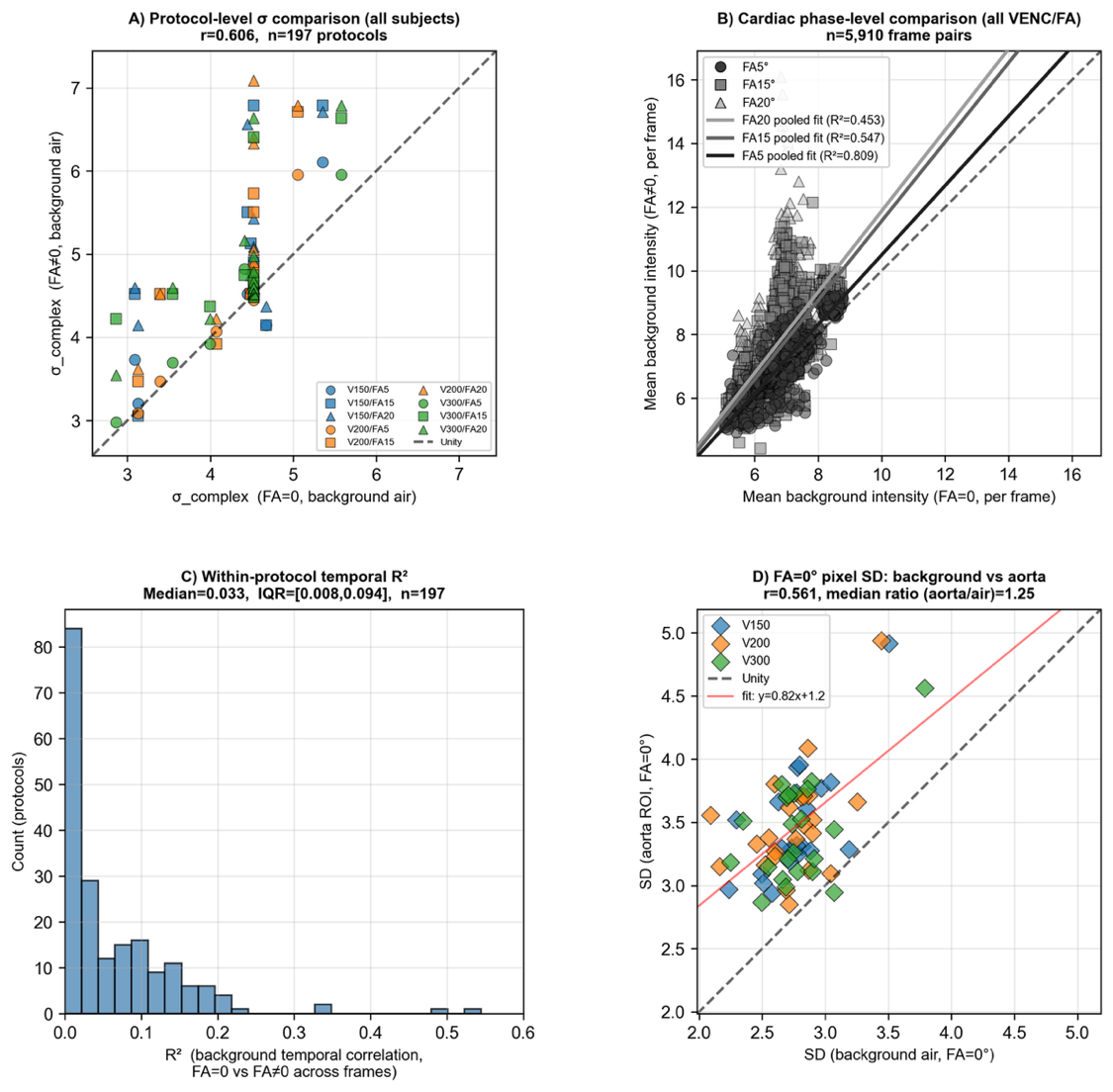

**Figure S1.** Validation of background noise estimation using FA=0° reference acquisitions. A) Protocol-level comparison of σ_complex_ estimated from background air in FA≠0° versus FA=0° acquisitions (r=0.606), demonstrating agreement in noise scale across acquisition conditions. B) Cardiac phase-level comparison of σ_complex_ estimates from background air in FA≠0° and FA=0° acquisitions (r=0.761), indicating consistent noise behavior within individual acquisitions. C) Distribution of within-protocol temporal stability of background noise across cardiac phases, quantified as Pearson r^2^ between FA=0° and FA≠0° background frame-mean time series (median r^2^=0.033, IQR: 0.008-0.094), demonstrating minimal frame-to-frame correlation and supporting temporal independence of noise. D) FA=0° noise quality-control analysis comparing background-derived σ_complex_ with the standard deviation of FA=0° magnitude signal within the aorta ROI (r=0.334, median ratio=0.66). This panel is interpreted as an order-of-magnitude QC check rather than a direct one-to-one comparison, as the aorta ROI contains low but nonzero signal and therefore does not follow the Rayleigh noise model assumed for background air.

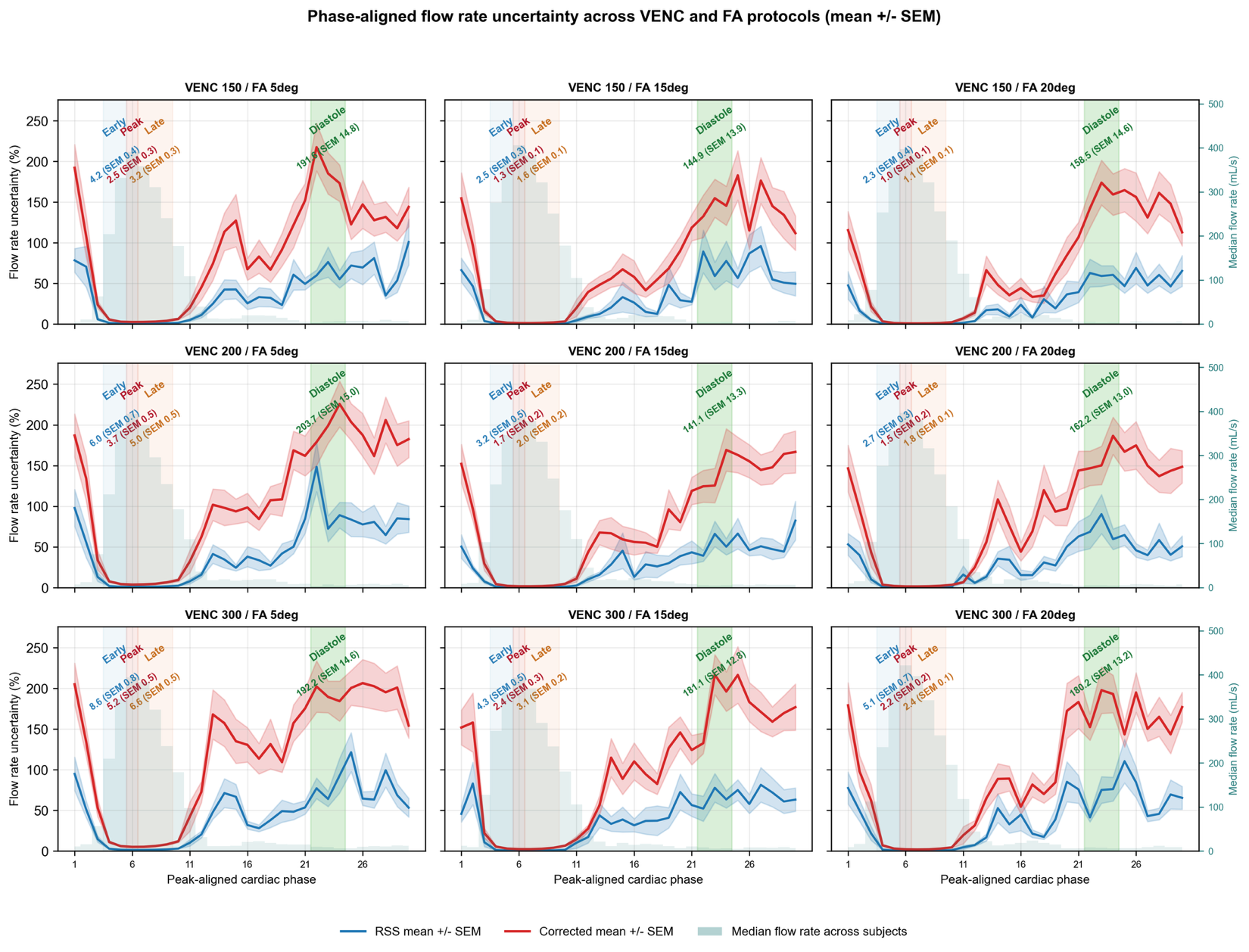

**Figure S2.** Phase-aligned instantaneous flow uncertainty across VENC and flip angle (FA) protocols. Phase-aligned cardiac cycle plots of instantaneous flow percent uncertainty (%unc) for all VENC (150, 200, 300 cm/s) and flip angle (FA=5°, 15°, 20°) combinations. Blue curves show mean %unc under the independent-voxel assumption (RSS), and red curves show mean %unc after spatial correlation correction; shaded regions represent ±SEM across subjects. Gray shading indicates the median absolute flow rate (|Q|) across subjects. Vertical shaded regions denote phase-based groupings relative to peak-aligned flow: early systole (blue), peak systole (red), late systole (orange), and diastole (green). Text annotations within each panel indicate mean %unc values (±SEM) for each phase bin. All curves are aligned such that peak systolic flow occurs at the same cardiac phase for each subject.

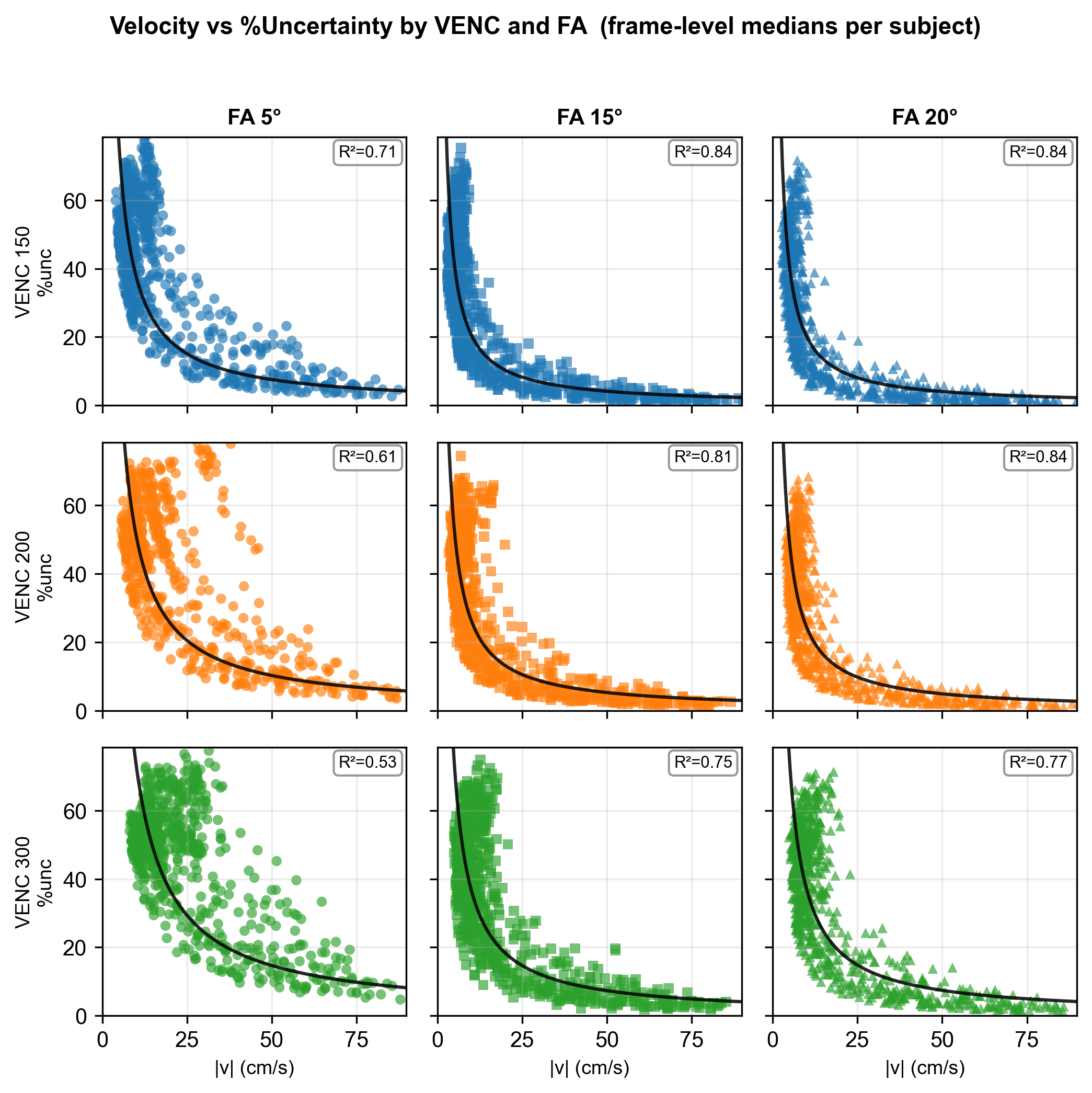

**Figure S3.** Protocol-specific relationships between velocity magnitude and percent uncertainty across VENC and flip angle combinations. Relationships between |v| and %unc are shown for each VENC (150, 200, and 300 cm/s) and FA (5°, 15°, and 20°) combination using frame-level aortic ROI median observations. Lower VENC and higher FA were associated with reduced uncertainty and tighter inverse relationships between velocity magnitude and %unc, with protocol-specific R^2^ values ranging from 0.53 to 0.84.
